# Health care workers’ internal bias toward men as HIV clients in Malawi and Mozambique: A qualitative study

**DOI:** 10.1101/2022.11.15.22282343

**Authors:** Kathryn Dovel, Rose Paneno, Kelvin Balakasi, Julie Hubbard, Amílcar Magaço, Khumbo Phiri, Thomas Coates, Morna Cornell

**Author notes:** **Corresponding Author:** Kathryn Dovel, Division of Infectious Diseases, David Geffen School of Medicine, 10833 Le Conte Ave, Los Angeles, CA 90095, /.

## Abstract

**Background:** Men are underrepresented in HIV services throughout sub-Saharan Africa. Little is known about health care worker (HCW) perceptions of men as clients, which may directly affect the quality of care provided, and HCWs’ buy-in for male-specific interventions.

**Methods:** Focus group discussions (FGDs) were conducted in 2016 with HCWs from 15 facilities across Malawi and Mozambique, and were originally conducted to evaluate barriers to universal treatment (not gender or internal bias). FGDs were conducted in local languages, recorded, translated to English, and transcribed. For this study, we focused on HCW perceptions of men as HIV clients, using inductive and deductive coding in Atlas.ti v.8, and analyzed codes using constant comparison methods.

**Findings:** 20 FGDs with 154 HCWs working in HIV treatment clinics were included. Median age was 30 years, 59% were female, and 43% were providers versus support staff. HCWs held strong implicit bias against men as clients. Most HCWs believed men could easily navigate HIV services due to their elevated position within society, regardless of facility-level barriers faced. Men were described in pejorative terms as ill-informed and difficult clients who were absent from health systems. Men were largely seen as “bad clients” due to assumptions about men’s ‘selfish’ and ‘prideful’ nature, resulting in little HCW sympathy for men’s poor use of care.

**Interpretation:** Our study highlights a strong implicit bias against men as HIV clients, even when gender and implicit bias were not the focus of data collection. As a result, HCWs may have little motivation to implement male-specific interventions or improve provider-patient interactions with men. Framing men as problematic places undue responsibility on individual men while minimizing institutional barriers that uniquely affect them. Implicit bias in local, national, and global discourses about men must be immediately addressed.

## INTRODUCTION

Engaging men in HIV services is critical to curbing HIV epidemics in sub-Saharan Africa. Yet efforts to engage men have been largely suboptimal,^1,2^ despite the rapid expansion of male-specific interventions. ^3,4^ Health care worker (HCW) perceptions of men as HIV clients may be a major barrier to men’s care,^5,6^ particularly since HCWs are the ones required to implement male-specific interventions.

HCWs may hold internal biases against men as clients. Men have historically been absent from global HIV guidelines and priorities,^7^ and when they are represented, they are often depicted as problematic, difficult clients or perpetrators of local epidemics.^8–10^ Such discourses can “trickle down” to local HCWs through HCW trainings and donor priorities.^11,12^ Further, HCWs often have limited exposure to men as clients due to how health systems are organized to prioritize women’s reproductive health and children’s health.^12^ In Malawi, national guidelines recommend that women of reproductive age attend a health facility 5-17 times a year for routine, non-acute services; men of the same age range are only recommended to attend once for HIV testing.^13^ Limited exposure to men as clients may limit HCWs’ comfort level interacting with men, and perpetuate broad stereotypes of men as clients.

HCW perceptions are thus critical to improving the HIV response for men. Negative perceptions and internal bias have been linked with poor quality care among men of color in the United States and the United Kingdom,^14^ and may have similar impact among men in sub-Saharan Africa, although to our knowledge this has not yet been studied. HCW perceptions also impact how male-specific interventions are implemented on the ground.^15–17^ According to the theory of Street-Level Bureaucracy (SLB), frontline service workers use extensive personal discretion to prioritize certain clients or interventions over others in the context of overwhelming workloads.^18^ Discretionary decisions are largely influenced by HCW perceptions of the clients they serve, particularly *who* is vulnerable and *why* particular populations do not succeed in care. Thus HCWs may prioritize clients perceived to be more vulnerable and worthy of focused care, while deprioritizing ‘difficult’ cases or clients deemed less “worthy.”^16^

A deeper understanding of HCW perceptions about men as HIV clients, *who* they see as most vulnerable to poor HIV-related outcomes, and *why* they believe men are missing from HIV services is needed to ensure effective implementation of HIV services for men. In this study, we apply constructs from SLB to qualitative data from HCWs in Malawi and Mozambique to understand HCW perceptions of men as clients.

## METHODS

We conducted secondary analysis on data from 20 focus group discussions (FGDs) with HCWs in Malawi and Mozambique (2016-2017). The original study aimed to understand provider perceptions of barriers to care under new universal treatment policies.^19,20^ This secondary study was conducted due to the unprompted, strong and morally charged perceptions HCWs reported about men as clients. The study included HCWs from 11 mid-level health facilities (6 in Malawi, 5 in Mozambique), with an average of two FGDs per facility. Facilities had medium to large ART cohorts, were in high HIV-burden areas, and were purposively selected by the study team to ensure representativity.

We conducted separate FGDs with ART providers and support staff (i.e., community health workers and HIV counselors). Focus group guides were developed based on existing literature on universal treatment and barriers to ART engagement. Guides included probes on differences by sex and age in these themes, but no specific questions about HCW perceptions of men as clients, vulnerability, or specific constructs from SLB. We used similar FGD guides in both countries, with questions slightly modified to ensure contextual relevance.

Two trained local research assistants led each FGD in the local language (Chichewa in Malawi and Portuguese as well as local languages including Changana and Macua, in Mozambique). FGDs lasted an average of 90 minute, and were audio recorded, translated, and transcribed into English by trained staff. We conducted regular spot checks of transcripts and translations to ensure completeness and accuracy.

For this secondary analysis, we developed a codebook based on existing literature on HCW perceptions of men in sub-Saharan Africa,^21^ internal bias literature,^14^ and relevant constructs from SLB regarding HCW perceptions of clients.^18^ RP and JH piloted the codebook on four FGD transcripts (two Malawi, two Mozambique), added iterative codes and clarified codebook definitions as needed. RM, JH, and KD reviewed the pilot coding and resolved discrepancies prior to finalizing the codebook. RP coded all remaining transcripts using the final codebook, allowing new themes to emerge using a modified grounded theory approach.^22^ KD, JH, RP, and KB met weekly to compare codes for consistency and resolve differences across FGDs. Coded data were analyzed within the overarching framework of SLB in Atlas.ti7.5.^23^

### Ethics

Ethical approval for the original trial was attained by the Institutional Review Board at University of California Los Angeles and the National Health Sciences Review Committee in Malawi. Participants were not paid for participation but were given refreshments during the FGDs. Oral consent was provided. No identifiers were collected.

## RESULTS

We analyzed 20 FGDs with 150 HCWs who actively supported the delivery of HIV testing and treatment services in Malawi and Mozambique: 12 FGDs in Malawi (101 respondents); eight FGDs in Mozambique (49 respondents). The median age was 30 years, 59% of HCWs were female, 43% were ART providers (versus support staff), and the median duration for a career in HIV service delivery was three years (see Table 1).

**Table 1.** Demographics of Health Care Worker respondents (n=150)

| Variable | Malawi (n=101) | Mozambique (n=49) | Total (n=150) |
| --- | --- | --- | --- |
| Age, median (IQR) | 31 (29-41) | 29 (28-39) | 30 (29-39) |
| Female | 70 (70%) | 19 (38%) | 59% |
| Occupation |  |  |  |
| Provider | 40 (40%) | 24 (49%) | 64 (43%) |
| Support Staff | 61 (60%) | 25 (51%) | 86 (57%) |
| Years working in HIV sector, median (IQR) | 3 (2-5) | 4 (2-7) | 3 (2-6) |

Two major themes emerged and are discussed below:

### Who is vulnerable to poor HIV-related outcomes: women are vulnerable, men have safety nets

Perceptions of vulnerability were frequently discussed in FGDs. HCWs often described systematic vulnerabilities experienced by women, while men were seen as having full agency to control their own health outcomes. HCWs recounted numerous stories of women being afraid to disclose their HIV-positive status for fear of losing their husband, or husbands taking the wives’ treatment themselves, making it impossible for women to adhere. To HCWs, societal gender inequalities and unequal power dynamics within relationships meant that men took advantage of women, leaving women unable to navigate HIV service utilization.

> *There are wives [taking ART] in hiding, and this influences her treatment adherence. Other wives do not reveal [their status] because they fear that the partner will beat them or they will get divorced. Nowadays, when the men find out [about their partners positive status], they abandon the family. (Mozambique, Support Staff)*
>
> *Most women fail to start ART because of challenges related to patriarchal dominancy. Most men out there still think in the outdated manner that they are superior than women and as such a woman is prevented from making an independent decision to start ART because she is afraid that she will risk breaking up her marriage. (Malawi, Support Staff)*

While HCWs readily described men’s poor use of health services, they believed men brought poor health “upon themselves”. Men’s increased power within the larger society led HCWs to believe men were responsible for their own choices. External factors were seen to play a minimal role in men’s ultimate use of HIV services.

> *While in men, the decision seems to be his own*… *Ahhh its easier for the men to start because the decision power he possesses. The woman, she understands. The woman understands the facts [about ART] very well, but the problem is in the power of decisions. (Mozambique, Support Staff)*

Some HCWs believed men were less motivated than women to seek HIV services because they had greater safety nets for health care if they became ill, making men less vulnerable to poor health outcomes. These HCWs argued that men could afford to avoid HIV services because they could rely on their wives and family to care for them. Women, however, were described as having minimal safety nets for themselves and their children, and therefore more concerned about seeking HIV services promptly since they were the only ones who would take care of themselves, and their family.

> *Women are always at home taking care of the family. They also care for their [own] health as well since there is no one to take care of their children when they are sick. But men do not really worry about their [own] health because they know that if they get sick then the woman will take of him and the children. (Malawi, ART Provider)*

### Why men are underrepresented in HIV services: individual men blamed for facility-level barriers

Perceptions of men’s elevated status in society led HCWs to believe that men had full agency over their own health seeking behavior. Thus, while HCWs readily identified facility-level barriers to men’s care, they also believed that men should be able to overcome these barriers. Men who did not overcome facility-level barriers were generally described as “too proud” or “selfish” to access care. Below are two examples of the disconnect between barriers identified and blaming individual men.

#### Men who do not overcome barriers for income-generation are too proud

HCWs recognized that time and travel requirements for accessing HIV services directly conflicted with men’s role as economic providers, a role HCWs believed men internalized as a core part of their identity.

> *Men are breadwinners and if a man comes here to collect ART and stumbles upon a long queue, they are easily discouraged to wait for an hour to get ART while they are still healthy. Instead, they start thinking of what they could accomplish in that one hour if they were to be working and so they choose to go back [home] and come back [to the facility] later when they are sick. (Malawi, ART Provider)*

However, few HCWs felt sympathetic toward the tension men experienced. Instead, they believed that most men could afford to forfeit income generation activities for monthly ART appointments, but that men chose work over HIV services due to pride and placing undue emphasis on earning money.

> *Pride! Men are so proud. They feel that they are so important. Maybe due to the fact that they usually fend for the family and that causes them to feel that they cannot afford to be sick, let alone go to the hospital on a monthly basis to collect ART. (Malawi, ART Provider)*
>
> *[Men are] Pompous! Pompous! Because they feel themselves to be of higher social status, so when they are told about [HIV], they don’t see the negative impacts right away and they don’t want these issues to disturb their normal [work]. (Malawi, ART Provider)*

HCWs believed men had too much power as the economic providers, and therefore would not listen to advice from their wives or HCWs. One HCW explained that since men are in control of the household, they assume the same power should extend to their health and health care.

> *They are different, men and women. Women do accept their counselling very quickly compared to men. Men they sometimes become troublesome to accept their results, so it can be difficult … They are head of the family and head of their immune system [laughter]. (Malawi, ART Provider)*

#### Men who do not overcome feminized health services are too selfish

HCWs frequently described how health facilities were tailored to women and children, creating unique barriers to men’s use of HIV services. Women were expected to attend health facilities and outreach clinics regularly for family planning, antenatal care, and children under-five years of age services, giving women extensive health information and multiple reasons to attend a health facility. In contrast, there was general consensus that men rarely attended health facilities – either for their own health or in support of others’ health. As a result, men were seen as having little exposure to health education and increased risk of unwanted HIV status disclosure when seeking HIV services – why would men attend a health facility expect for HIV services?

> *Because of the responsibilities that we [women] have – like falling pregnant, coming for antenatal care, delivering the baby and looking after it – we spend more time at the hospitals than men, and there is no way that you can be at a hospital and leave without getting some of the crucial information that you have to know at that time. Unlike men, they shun away from [services]. (Malawi, ART Provider)*
>
> *… A lot of people haven’t realized yet that this disease [HIV] is like any other. Sometimes men fear meeting people they know here at the health facility, and that makes them stay home [not start ART]. (Mozambique, Support staff)*

While the organization of health services was widely recognized as a barrier to men’s care, men who were unable or unwilling to access care through feminized health services were blamed for their poor engagement. HCWs used morally-charged language, describing these men as “irresponsible” or “too selfish” to face the additional challenges associated with female-focused services.

> *Men are usually self-centered and therefore do not see anything wrong with not taking ART, while women think about their families, “What will happen to my family if I fall sick?” … men are too selfish, such that they don’t care even when they are HIV-positive. (Malawi, Support staff)*
>
> *Men need punishment. If man comes to the health unit it is because he is [ill] … [Men] are confusing. I think men believe that because he is a man, he can never get sick. He does not want to show his weakness. So that’s why it takes him to the hospital unit [he becomes very sick]. (Mozambique, Provider)*

While HCWs recognized that men risked unwanted disclosure by simply attending an ART clinic, this did not elicit sympathy because many HCWs believed men’s primary reason for avoiding disclosure was to avoid being seen as weak or “risky sexual partners”. Men’s fear of disclosure was thus seen as selfishly motivated, and not based on concern for their families.

> *Provider 1: They [men] fear to be rejected by society, to lose their wives. That’s why they do the HIV test in another health center …*
>
> *Provider 2: Most men have this conception, maybe if his neighbor sees him going to hospital he will know that he’s in that position [HIV-positive], but women don’t resist much. (Mozambique, Support Staff)*

Overall, HCW’s described how individual men were expected to overcome extensive facility-based barriers to care, and men’s character traits of pride and selfishness were believed to make it difficult for them to do so.

## DISCUSSION

Our study highlights a strong implicit bias against men held by both male and female HCWs in Malawi and Mozambique, even when gender and implicit bias were not the focus of the data collection process. HCWs saw women as vulnerable to poor HIV-related outcomes due to societal inequalities, and unable to overcome structural barriers to care. In contrast, most HCWs believed men could easily navigate HIV services due to their elevated position within society. Men were described in pejorative terms as ill-informed and difficult clients who were largely absent from the health care system. While HCWs recognized facility-level barriers to care for men, they had little sympathy for men, largely seeing men as “bad clients” due to assumptions about men’s ‘selfish’ and ‘prideful’ character.

HCW perceptions of men were often in direct conflict with existing literature, speaking to the disconnect between stereotypes and reality. HCWs largely described men as controlling and uncaring toward their female partners, uninterested in their own health or the health of their families, and largely absent from health facilities. Yet such stereotypes are now being refuted throughout the region. In Kenya and Tanzania, over 80% of women living with HIV reported positive male partner reactions to disclosure of their HIV status, and an overwhelming majority reported partner support for HIV service utilization^24,25^ although fear of IPV is an established barrier to women’s care.^26^ Other studies show that men are invested in their children’s well-being^27^ and are motivated by educational and economic attainment for their families.^28^ Notably, in a community-representative survey in Malawi, 80% of men had visited a health facility within the past year, with nearly half attending facilities as caregivers for family members or friends.^29^ These finding are in stark contrast to HCW perceptions of men as so self-absorbed that they did not care about the consequences of not accessing health services. In the USA and UK, harmful stereotypes of clients from minority groups may be used by HCWs to justify poor health outcomes and remove responsibility from HCWs or health institutions.^30,31^ Our data suggests a similar phenomenon may be taking place for heterosexual African men within the local HIV epidemics in Malawi and Mozambique. Such stereotypes deserve immediate attention as they may negatively impact HCW buy-in and implementation of male-specific interventions,^16,18^ and negatively impact patient-provider interactions.^14^

HCWs expected men to overcome structural barriers to HIV services that were not expected of women. Men’s generally elevated status in society meant that HCWs could not see men as vulnerable in other contexts, such as health service utilization. Men were believed to have enough agency to navigate their own health care, despite the numerous barriers faced. This attitude is not unique to HCWs. Vulnerability within global health is often used to describe women and children exclusively, regardless of the specific context.^10,32^ For instance, in a recent review of 201 prominent global health organizations, only 42% had gender-responsive strategies to meet the needs of both women and men. Not one primarily targeted the health of men, even though men had worse outcomes than women in all the top ten contributors to global morbidity and mortality.^33^ Global health must embrace more nuanced, intersectional definitions of vulnerability which acknowledge that groups who are powerful in one social sphere may be vulnerable in another.^34^ Men face important differences in agency based on their intersectionality of identities,^35,36^ and the structure of health systems in which they are asked to engage.^12^

Negative perceptions of men as clients should not come as a surprise. Such perceptions fit neatly within the historical global discourse of men within African HIV epidemics.^8–10^ Multilateral agency and donor discourses have tended to paint men as the problem and difficult clients. While there are now positive shifts in how men are represented in HIV responses,^37^ donor priorities and HCW trainings over the years have lasting effects on HCW perceptions. We found similar perceptions among HCWs in both Malawi and Mozambique, in different cultural and language contexts, and among male and female HCWs. Such similarities may speak to the powerful influence of multilateral bodies and funding agencies on local perceptions and lived experiences within HIV services,^38^ Our findings also highlight the role of harmful gender norms in shaping health institutions and patient-provider interactions. Perceptions of men as a homogenous group that is strong, invincible, and with unlimited agency are key examples of HCWs internalizing harmful stereotypes regarding what is normative masculinity and how men should behave.^12,39^

What can be done? There is an urgent need to change global HIV discourse about men – shifting from blaming men to understanding and addressing the structural barriers men face. HIV priorities and policies of multilateral partners and donors require urgent review to reflect the complex realities faced by men and remove morally charged language about the men we are now trying to reach. Additionally, local efforts are needed to change HCW perceptions. Potential strategies include sensitization workshops for HCWs to de-stigmatize men as “bad clients”, like those used for Welcome Back campaigns^40^ and stigmatized populations. Further, facility-level barriers should be addressed so that individual men are not responsible for overcoming structural barriers to care. Finally, it is time to recognize men’s unique vulnerability to poor service utilization and to prioritize research on HIV in heterosexual men in SSA, as well as other marginalized populations who may not frequent facilities through reproductive or children’s health services.

Several limitations should be noted. First, this was a secondary analysis and data were not collected with HCW perceptions of men in mind, potentially limiting the depth of analysis. However, this could also be regarded as a strength, in that such strongly gendered stereotypes and moral characterizing of men emerged in secondary analysis of data in different contexts without prompting. Second, we are unable to differentiate between perceptions from male and female HCWs. Finally, we are unable to assess how HCW internal bias affects the quality of services. Additional research is needed on *how* such perceptions affect HCW interactions with male clients and the implementation of male-focused strategies, as well as the most effective strategies to change these negative perceptions.

## CONCLUSION

Our study highlights the strong negative perceptions HCWs hold toward men as clients, even when implicit bias was not the focus of the data collection process. Framing men as problematic clients places undue responsibility on individual men, while minimizing health system barriers that uniquely affect them. Global, national, and local guidelines should be reviewed to minimize negative stereotypes against men. Interventions targeting men should be paired with HCW sensitization strategies to address implicit bias toward men as clients.

## Data Availability

The data that support the findings of this study are available from the corresponding author, KD, upon reasonable request.

## Financial disclosure

This work was funded by the Bill and Melinda Gates Foundation (INV-001423), the National Institute of Mental Health (NIMH R01-MH122308), Fogarty International (K01-TW011484-01) and UCLA GSTTP.

## Declaration of interests

We declare no competing interests.

## Authors Contributions

KD and RP conceptualized the study. KD is responsible for funding acquisition. KD, AM, and KP developed the parent study protocol and materials.. KD and RP developed the formal analysis plan and RP and JH analyzed the data. KD and RP wrote the original draft and JH, KB, KP, AM, TC, and MC edited following drafts. All authors have read and approved the final manuscript.

## Notes

### Competing Interest Statement

The authors have declared no competing interest.

### Clinical Trial

None

### Author Declarations

Ethical approval for the parent study was attained by the Institutional Review Board at University of California Los Angeles and the National Health Sciences Review Committee in Malawi. Participants were not paid for participation but were given refreshments during the FGDs. Oral consent was provided. No identifiers were collected.

